# PREVALENCE AND ASSOCIATED RISK FACTORS OF SCABIES AMONG UNDER-FIVE-YEAR-OLD CHILDREN IN MAFINGA DISTRICT, ZAMBIA

**DOI:** 10.1101/2025.05.15.25327682

**Authors:** Michael Kay Ndhlovu, Lungowe Sitali-Zimba

## Abstract

**Background:** Scabies remains a significant public health concern in sub-Saharan Africa, particularly among children and the elderly. It is mostly found in developing countries, and it has health effects due to its discomfort and secondary bacterial infection.

In Zambia, there is limited data on the burden and determinants of scabies, particularly in high-risk areas like Zumbe Health Post (HP) in Mafinga District. Therefore, the purpose of this study was to determine the prevalence of scabies and its associated risk factors among children under five years old in Mafinga District, Zambia.

**Methodology:** A cross-sectional study design was employed. Two sampling techniques were used: purposive and simple random to select 307 under-five children and their caregivers. Scabies diagnosis was based on clinical examination set by the International Alliance for the Control of Scabies (IACS, 2020), and data on associated risk factors was collected via structured questionnaire. Univariable and multivariable regression analyses were conducted to identify associations between scabies and risk factors.

**Results:** The prevalence of scabies was 5.21% (16/307; 95% CI: 2.8%–7.6%). Most cases (68.75%) were mild. Multivariable analysis identified that caregivers who took longer to fetch water had significantly higher odds of scabies among their children (AOR: 3.8; 95% CI: 2.61–5.82; p< 0.001). Higher caregiver education levels were associated with lower scabies risk (AORs: 0.02– 0.03), and high knowledge of scabies among caregivers was significantly associated with lower scabies prevalence (p = 0.02).

**Conclusion:** Scabies prevalence in Zumbe Health Post was moderate. This Moderate highlights the need for targeted interventions in the community. Efforts should focus on improving caregiver education and access to clean water to help reduce the incidence of this condition among children. Provision of health education and improvement of hygiene practices are suggested. These strategies can empower caregivers with the knowledge and resources necessary to prevent scabies transmission.

**Author summary:** This study was carried out in the Zumbe catchment area in Mafinga district, Zambia. It aimed to assess the prevalence of scabies and associated risk factors among under five-year-old children. The study involved guided questionnaire and clinical skin examination for scabies. Finding revealed moderate prevalence of Scabies and access to water on time taken and knowledge level as associated risk factors of scabies. There is a need to improve on hygiene practice and community health promotion to protect the children from Scabies.

## Introduction

Scabies is defined by (Heukelbach and Feldmeier, 2006) as a “an ectoparasitic infestation caused by the mite *Sarcoptes scabiei var hominis* which is an obligate parasite that completes its entire life cycle on humans. Female mites burrow into the skin and lay eggs, eventually triggering a host immune response that leads to intense itching and rash”.

It is a neglected but significant public health challenge globally (Engelman et al., 2019). Despite being preventable and treatable, it affects over 200 million people worldwide, with the highest burden in developing countries (WHO, 2020).

The world prevalence of scabies ranges from 0.2% to 71% and mostly affects children and the elderly (Hay et al., 2014; Engelman et al., 2019). In Africa, scabies prevalence ranges from as low as 3.8% to as high as 65%, depending on the area and the economy of the country (Sherbiny et al., 2017; Ugbomoiko et al., 2018). High prevalence is related to overcrowding, poor hygiene, and limited water access (Ibadurrahmi et al., 2016; Azene et al., 2020). Children are most affected due to low immune systems and dependence on caregiver hygiene (Romani et al., 2015; Matthews et al., 2021).

In Africa, prevalence rates of 5–35% have been reported, particularly among school-age children during the wet season (Chosidow, 2006; Yeshanew et al., 2022).

The main effect of scabies is generalized itching due to the host immune response against the mites which may disturb sleep, school and life in general and the secondary effects are opportunistic bacterial infection likes Impetigo caused by *Staphylococcus aureus* and *Streptococcus pyogenes* taking advantages of persistent itching due to scabies (Engelman et al., 2013.

Scabies risk factors globally recognized can be summarized in to: poor hygiene practices, which are consistently linked with increased scabies transmission (Heukelbach and Feldmeier, 2006); caregiver knowledge which are causes, transmission, symptoms, and prevention which are critical to early detection and effective management (Amoako et al., 2020; Nanyunja et al., 2020); and water significantly influences scabies transmission (WHO, 2020). The risk factors of scabies in Zambia remain underexplored (Simuyemba, 2020).

In Zambia, data on scabies are limited, with some outbreaks in institutional settings and no national strategy to prevent scabies (Mwanza et al., 2018; Zambia MOH, 2021). This study aims to address these gaps by focusing on children under five in the Zumbe Health Post catchment. Findings will help develop local intervention. No studies have been done on scabies in Mafinga District, Muchinga Province, Zambia.

## Methods

### Study area

The study was conducted in Zumbe Health Post, Mafinga District, which is 170km from Chasili, the provincial headquarters of Muchinga Province, Zambia.

### Study design

A cross-sectional study design was used to determine the prevalence of scabies and associated risk factors among under-five-year-old children in Zumbe HP.

### Sample size and sampling procedure

Two sampling techniques were used in this study. Purposefully sampling to select the study area because Zumbe Health Post recorded the highest cases of scabies in Mafinga District (Mafinga DHO, 2023) and simple random sampling by entering the population of 1,333 under-five-year-old children (Central Statistics Office, 2023) from the cohort register of Zumbe Health Post into an Excel sheet and carrying out a simple random sample of 307 under-five-year-old children and their caregivers based on the sample size calculation. The under-five-year-old children and their caregivers sampled were seen in one week during the facility outreach program.

### Data collection tools and techniques

Scabies was screened clinically using physical examination outlined by IACS (2018) guidelines by experienced clinical officers. The lesions were observed during the physical examination on the hand, wrist, arm, elbow, axilla, leg, foot, abdomen, thorax, mammilla area, back, buttock, genital/inguinal area, and head.

A schedule (guided questionnaire) was administered to under-five children’s caregivers based on (Collinson et al., 2020; Kouotou et al., 2016a; Azene et al., 2020) and it contains four parts, which include information on socio-demographic factors, hygiene practice, knowledge levels, and access to water. It was then translated from English to Bemba, the local language in use. The schedule was collected by two community health assistants and one environmental health officer who was supervising the data collectors.

### Data analysis and processing

The data were entered using Microsoft version 16 and analyzed using the SPSS statistical package for Windows, version 16.0. Descriptive statistics of texts and tables were used. Factors associated with scabies were identified using multivariable logistic regression analysis, which was fitted, and the corresponding adjusted odds ratio (AOR) and 95% CI were used. Significant results were characterized statistically by a p-value < 0.05. The Hosmer-Lemeshow goodness-of-fit statistic was used to assess whether the necessary assumptions for the application of multiple logistic regression are fulfilled. For this study, a p-value was found to be 0.77, implying that the model was a good fit.

### Ethical considerations

The study was approved by the University of Zambia Biomedical Research Ethics Committee (UNZABREC) (Ref No: UNZA-4204/2023) and National Health Research Agency (NHRA) (Protocol No: NHRA0006/07/09/2023). Permission to conduct the study was sought from Muchinga Provincial Health Office and Mafinga Distict Health Office. A written informed consent is obtained from the caregivers before they participated in the study. We explained the study object, autonomy, and confidentiality of the questionnaire, and the right to participate and withdraw at any time to both caregivers and participating children before and during collecting data. Children who hard positive clinical scabies were treated.

## Results

As shown in Table: 1 a total of 307 Under-five-year-old children with their caregivers 307 were selected in this study with a response rate of 100%. The age of under-five was categorized into four categories. Under 2 years’ (121) (39.4%), 2-3 years’ 94(30.6%), 3-4 years’ 57(18.6%) and 4-5 years’ 57(18.6%). The scabies cases were highest among under 2 years’ and 2-3 years’ (6) (37.5%) followed by 3-4 years’ and 4-5 years’ (2) (12.5%). Although there was no statistical evidence of association between age and scabies cases (p-value = 0.09).

**Table 1.**
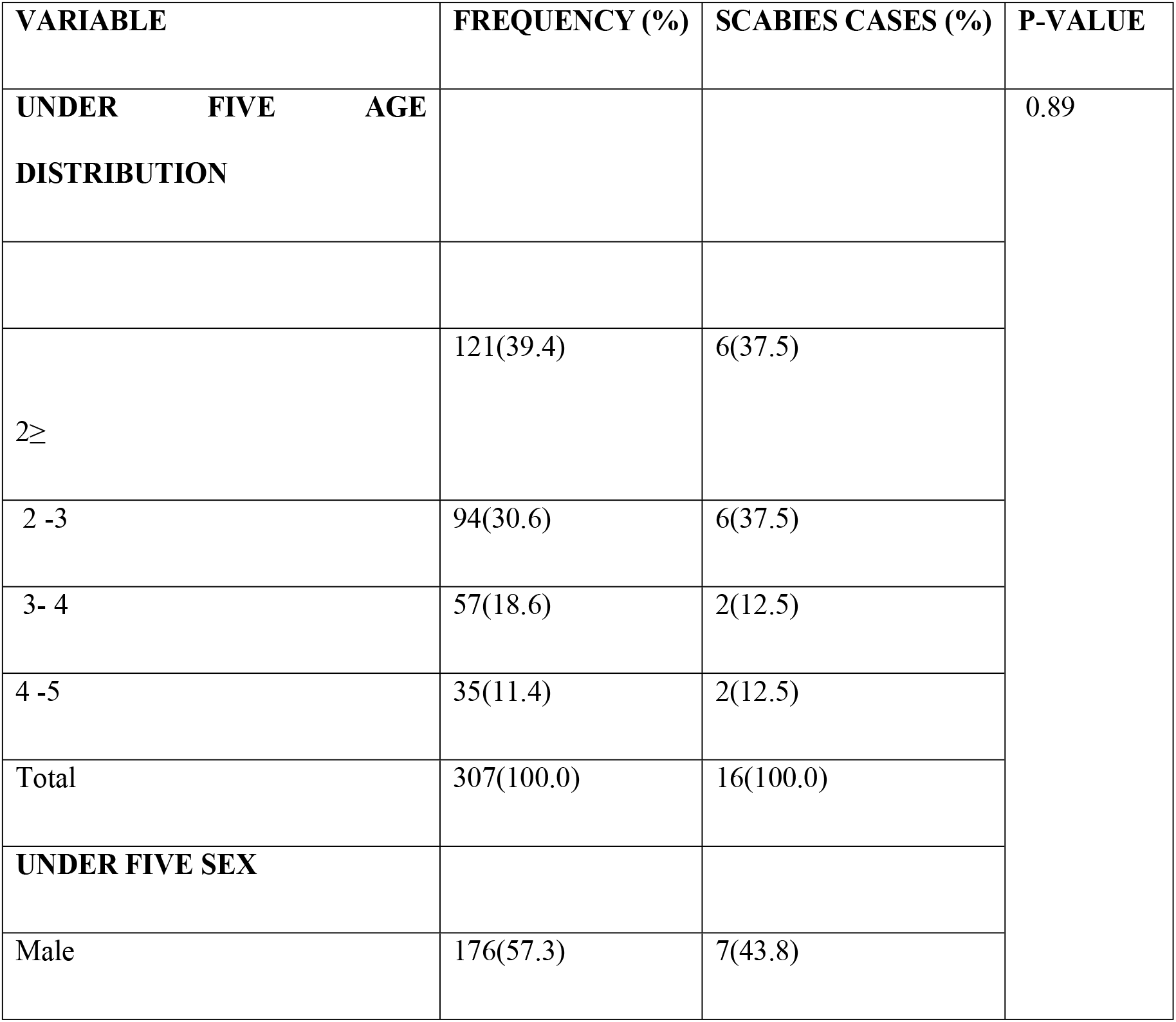
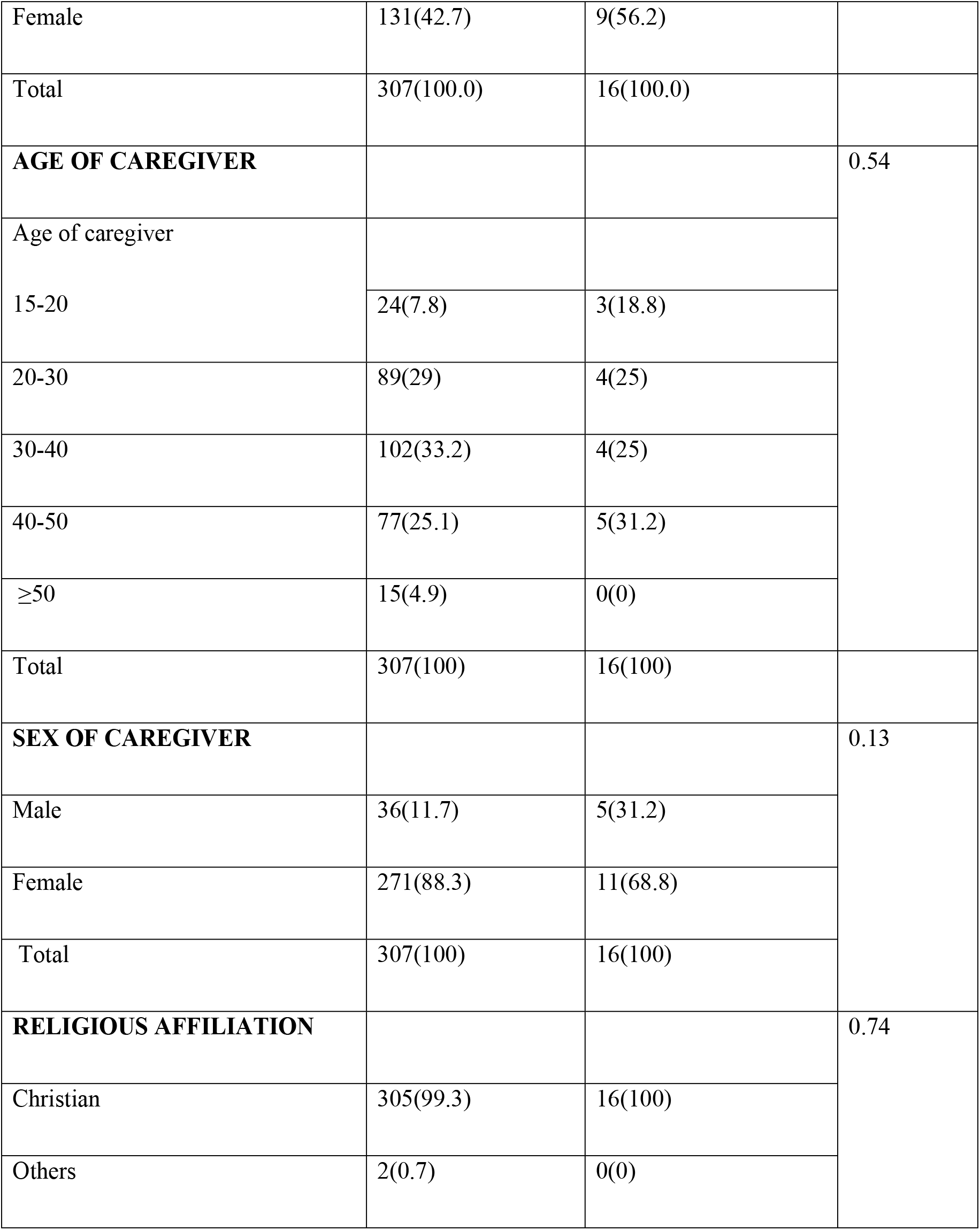
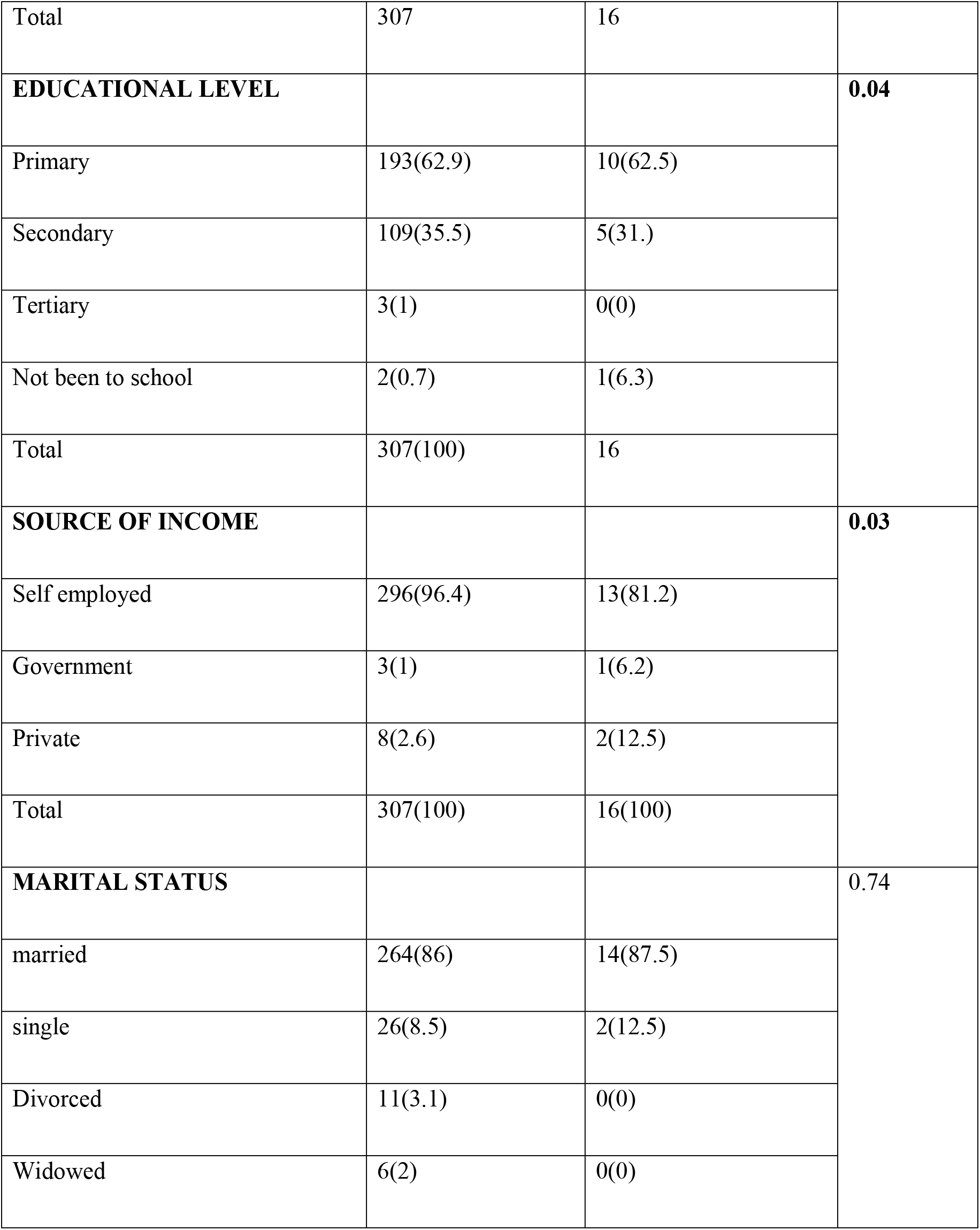
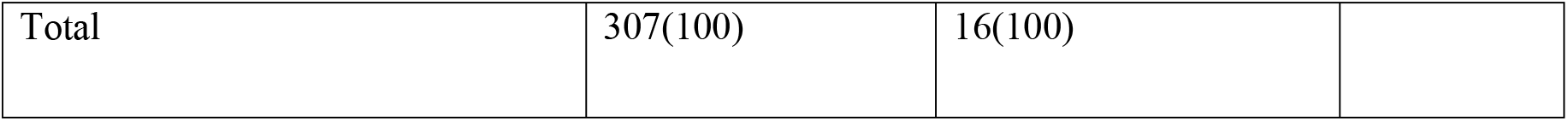
Social demographic characteristics of the study population.

### Prevalence of scabies

Results obtained from scabies screening of 307 Under-five-year-old children showed 16(5.2%) clinical positive cases and 291(94.8%) clinical negative cases. Thus the prevalence of scabies among the Under-five-year-old children was 5.21% (95% CI 7.6%, 2.8%). Out of the clinical positive cases cases11/16(68.75%) had mild, 4/16(25%) had moderate, and 1/16(6.25%) had severe lesion. This is shown in table 2.

**Table 2.** Scabies screening.

| VARIABLE | FREQUENCY (%) | CI |
| --- | --- | --- |
| **Scabies screening** |  |  |
| Clinical positive case | 16(5.2) | 7.6-2.8 |
| Clinical negative case | 291(94.8) |  |
| Total | 307 (100) |  |
| **Severity of illness** |  |  |
| mild | 11(3) | 3.6,2.4 |
| moderate | 4(1.3) | 2.3,0.3 |
| severe | 1(0.3) | 0.5,0.1 |
| Total | 16(100) |  |

**Table 3:**
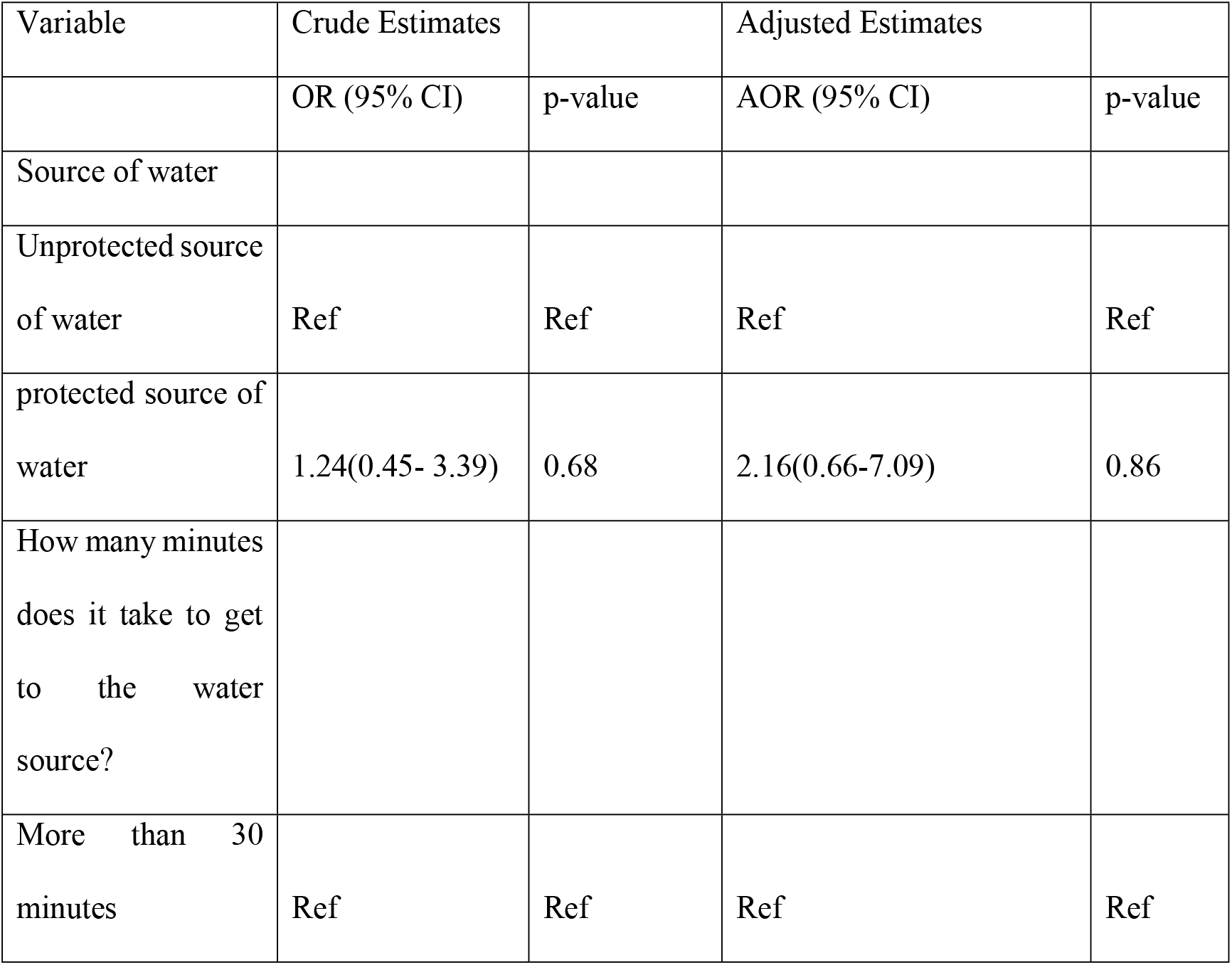
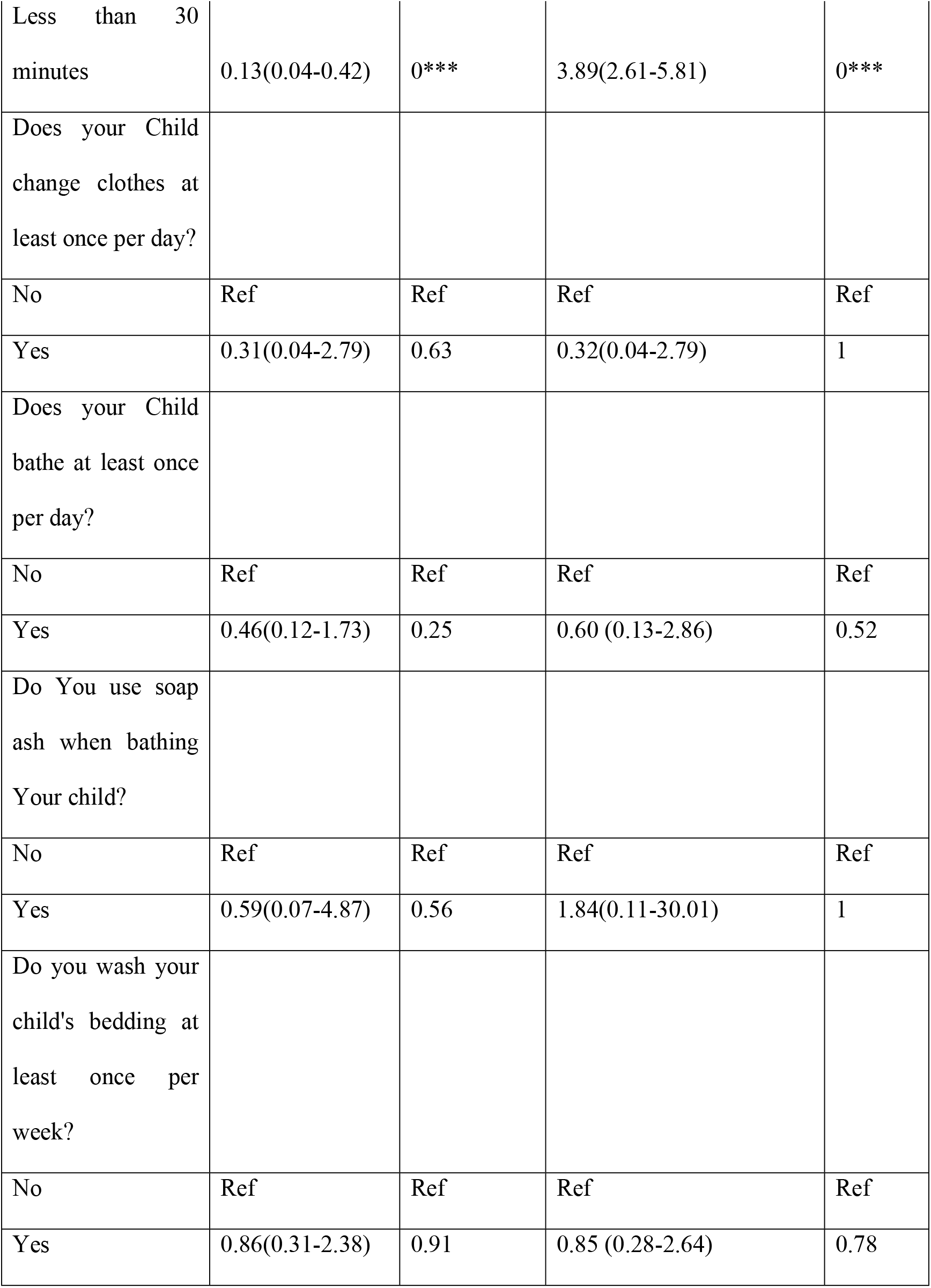
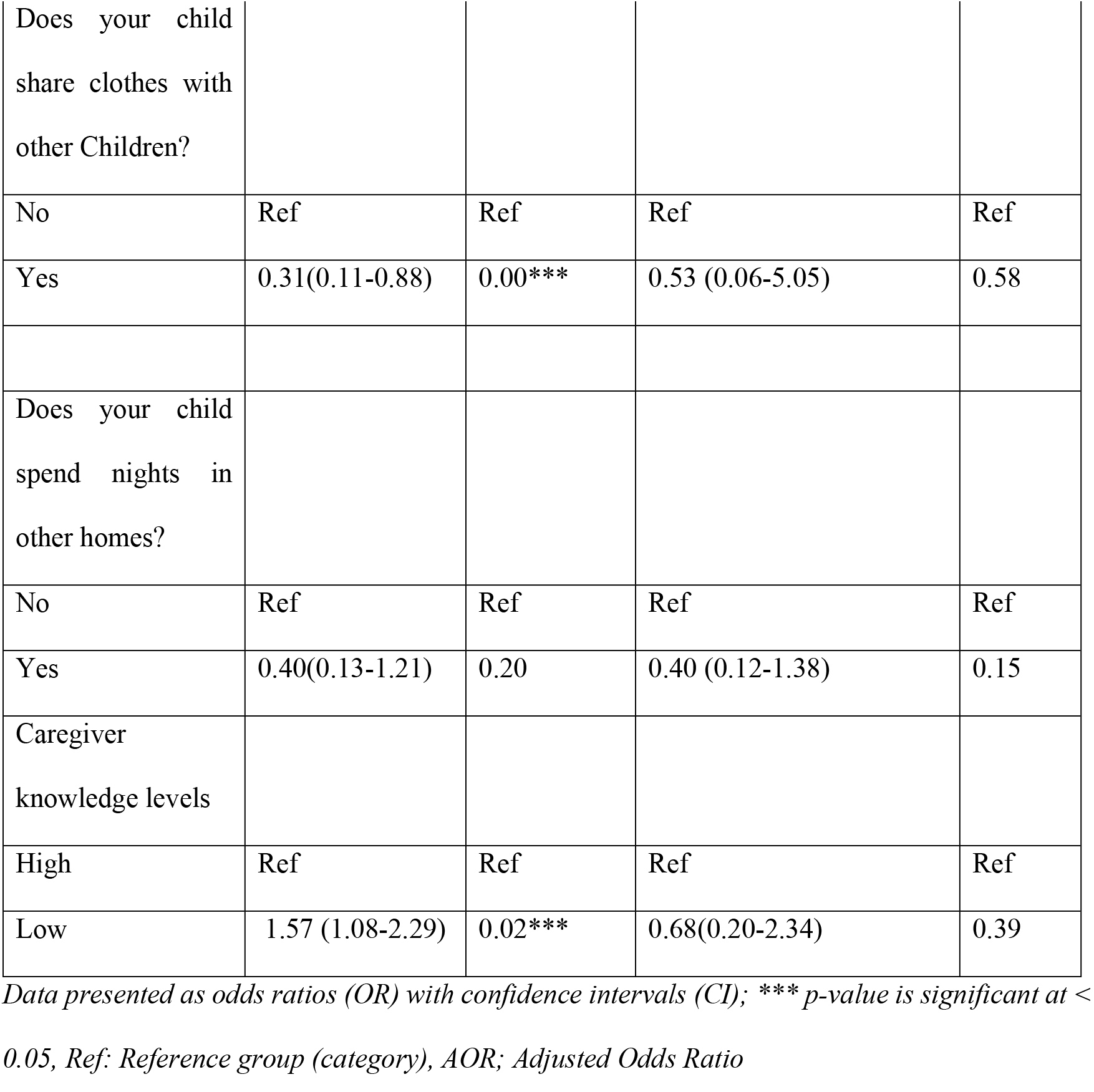
Univariate and multivariate logistic regression.

| Variable | Crude Estimates |  | Adjusted Estimates |  |
| --- | --- | --- | --- | --- |
|  | OR (95% CI) | p-value | AOR (95% CI) | p-value |
| Source of water |  |  |  |  |
| Unprotected source of water | Ref | Ref | Ref | Ref |
| protected source of water | 1.24(0.45- 3.39) | 0.68 | 2.16(0.66-7.09) | 0.86 |
| How many minutes does it take to get to the water source? |  |  |  |  |
| More than 30 minutes | Ref | Ref | Ref | Ref |
| Less than 30 minutes | 0.13(0.04-0.42) | 0*** | 3.89(2.61-5.81) | 0*** |
| Does your Child change clothes at least once per day? |  |  |  |  |
| No | Ref | Ref | Ref | Ref |
| Yes | 0.31(0.04-2.79) | 0.63 | 0.32(0.04-2.79) | 1 |
| Does your Child bathe at least once per day? |  |  |  |  |
| No | Ref | Ref | Ref | Ref |
| Yes | 0.46(0.12-1.73) | 0.25 | 0.60 (0.13-2.86) | 0.52 |
| Do You use soap ash when bathing Your child? |  |  |  |  |
| No | Ref | Ref | Ref | Ref |
| Yes | 0.59(0.07-4.87) | 0.56 | 1.84(0.11-30.01) | 1 |
| Do you wash your child's bedding at least once per week? |  |  |  |  |
| No | Ref | Ref | Ref | Ref |
| Yes | 0.86(0.31-2.38) | 0.91 | 0.85 (0.28-2.64) | 0.78 |
| Does your child share clothes with other Children? |  |  |  |  |
| No | Ref | Ref | Ref | Ref |
| Yes | 0.31(0.11-0.88) | 0.00*** | 0.53 (0.06-5.05) | 0.58 |
| Does your child spend nights in other homes? |  |  |  |  |
| No | Ref | Ref | Ref | Ref |
| Yes | 0.40(0.13-1.21) | 0.20 | 0.40 (0.12-1.38) | 0.15 |
| Caregiver knowledge levels |  |  |  |  |
| High | Ref | Ref | Ref | Ref |
| Low | 1.57 (1.08-2.29) | 0.02*** | 0.68(0.20-2.34) | 0.39 |

*Data presented as odds ratios (OR) with confidence intervals (CI); \*\*\* p-value is significant at < 0.05, Ref: Reference group (category), AOR; Adjusted Odds Ratio*

### Factors associated with scabies

In univariable analysis, under five age, under five sex, age of caregiver, sex of caregiver, religious affiliation, educational level secondary, marital status divorced and widowed, source of water, caregiver’s knowledge levels of scabies were significantly associated with the presence of mites. on the other hand, source of income, marital status married, time taken to water source, changing child’s clothes at least once per day, bathing the child at least once per day, use of soap ash when bathing a child, washing child’s bedding at least once per week, child sharing clothes with other children, child spending nights in other homes appeared as protective factors.

After multivariable analysis, time taken to water source (adjusted or (AOR) 3.8, 95 % CI: 2.61-5.82; P > 0.00), was the independent factor associated with scabies and educational level primary (adjusted or (AOR) 0.02, 95 %CI:0. 0,0.57; P >0.02, secondary (adjusted or (AOR) 0.03, 95 % CI: 0.00,0.81; P > 0.04 was a protective factor.

## Discussion

The present study aimed to assess the prevalence of scabies and its associated risk factors, specifically hygiene practices, caregiver knowledge, and access to water, among under-five-year-old children in the Zumbe Health Post catchment area of Zambia. The prevalence of scabies found in this study (5.21%) is below the WHO scabies alert threshold of 10% (World Health Organization, 2019). However, it aligns closely with the 4.7% prevalence reported in Ibanda, Nigeria (Ogunbiyi *et al*., 2005), and is higher than the 2.9% prevalence found in another study in Nigeria (Sambo *et al*., 2012) and the 4.1% prevalence recorded in Ethiopia (Azene *et al*., 2020). However, high prevalence has been reported, ranging from 10% to 71% in Nigeria (Anderson and Strowd, 2017), Gambia’s Sukuta region (Armitage *et al*., 2019), and Fiji (Steer *et al*., 2009). The difference in prevalence rates across different studies can be due to many reasons, such as family size, as found by Azene *et al*. (2020), in which large family size increased the chances of scabies, and the availability of hand-washing materials, as concluded by Kouotou *et al*. (2016), which may prevent scabies due to clean water or soap access.

This study identified that children who did not share clothes with others had a lower risk of scabies, which agrees with studies conducted in Indonesia, Cameroon, and Ethiopia (Mallongi *et al*., 2018; Kouotou *et al*., 2016a; Ejigu *et al*., 2019b). Personal hygiene is key in preventing and controlling scabies. Knowledge was associated with scabies prevalence in this study. Most caregivers had high knowledge levels, which could have contributed to the moderate prevalence of scabies because it could lead to early treatment, detection, and improved hygiene practices (Tresnasari *et al*., 2019; Liu *et al*., 2022; Lapeere *et al*., 2005). However, a study conducted in Indonesia concluded that knowledge alone cannot prevent scabies if other factors like low access to water and poor hygiene practices remain (Tresnasari *et al*., 2019).

## Conclusion

It was found that access to a protected water source was related to scabies in this study. This agrees with findings from Ejigu *et al*. (2019a) and Amoako *et al*. (2023). Nevertheless, access to protected water sources does not guarantee scabies prevention; it must be combined with improved hygiene practices such as regular bathing and washing of clothes, which are essential for disease control (Ararsa *et al*., 2023; Ejigu *et al*., 2019a; Amoako *et al*., 2023; Ayele *et al*., 2023). The moderate scabies prevalence observed in this study may be credited to a combination of protective behaviours, high caregiver awareness, and improved water access. These interconnected factors emphasize the importance of a multifaceted public health approach that integrates hygiene promotion, education, and infrastructure improvements to effectively reduce scabies transmission.

## Limitations of the Study

This study has several limitations that should be acknowledged: the cross-sectional study design does not establish causal relationships, and clinical examination of scabies may be prone to misdiagnosis. Additionally, confirmation through either dermoscopy or microscopy is necessary. Accessing the sample at a health facility like Mafinga District presents limitations, potentially leading to selection bias, as only individuals with more severe symptoms are likely to seek treatment. Furthermore, the hard scabies outbreak in Mafinga District a year prior could influence the results due to the interventions that were implemented

## Data Availability

All relevant data are within the manuscript and its Supporting Information files. Additional data may be made available from the corresponding author upon reasonable request.

## Acknowledgments

We would like to thank the Ministry of Health Zambia under Mafinga District Health Office for affording the team the opportunity to carry out the investigation. We are equally thankful to the Zumbe communities and participants who contributed to this study. Our gratitude ias also extended to the facility staffs in Zumbe for helping with data collection and navigating the catchment area.

## Authors’ contributions

All the authors actively participated during the conception of the research issue, development of a research proposal, interpretation, and writing various parts of the research report. Ndhlovu did data collection, analysis and prepared the manuscript and Sitali supervised the write up. All the authors read and approved the final manuscript.

## Funding

There was no funding for this research, but Mafinga District Health Office assisted with transport.

## Notes

### Competing Interest Statement

The authors have declared no competing interest.

### Funding Statement

The author(s) received no specific funding for this work.

